# Single-cell eQTL mapping identifies disease-specific genetic control of COVID-19 infection

**DOI:** 10.1101/2025.06.30.25330463

**Authors:** Jennifer Astley, Andrew Kwok, Benjamin Hollis, COvid-19 Multi-omics Blood ATlas (COMBAT) Consortium, Calliope A. Dendrou, Steve Sansom, Julian C. Knight, Alexander J Mentzer, Yang Luo, Luke Jostins-Dean

## Abstract

The impact of genotype on gene expression can depend on both cellular and organismal context. Here, we leverage an extensive blood atlas of genotyped patients with varying severity of infection produced by the COVID-19 Multi-omics Blood ATlas (COMBAT) Consortium to study the role of genetic regulation on gene expression in a context-specific manner.

We analyzed single-cell transcriptomic and genome-wide genetic data from ∼500,000 cells and 76 donors of European ancestry. Across 15 cell types, we identified 2,607 independent cis-eQTLs in high linkage disequilibrium (R2>0.8) with 48 infectious and 386 inflammatory disease-associated risk variants, including rheumatoid arthritis (RA) and inflammatory bowel disease (IBD). Notably, we found infection-specific eQTLs absent from a general population dataset (OneK1K), such as *REL*, *IRF5* and *TRAF*, all of which were differentially regulated by infection and whose variants are associated with RA and/or IBD. We also identified infection-modified eQTLs, including *RPS26* and *ADAM10*, implying that the regulatory sequences context of these genes may play a role in specific immune cell subsets in infection.

Our work demonstrates that the overriding effect of genetics on gene expression in blood immune cells is independent of infection status or severity. However, small numbers of eQTLs are modified by infection, and these differences can illustrate potentially important immune biology.

## Introduction

Large-scale studies of the genetics of gene expression, designed to map expression quantitative trait loci (eQTLs), have become one of the major tools used to investigate the mechanistic role of disease-associated risk variants (Aguet et al., 2020; Kerimov et al., 2021). This is particularly true for studying variants associated with risk of infectious or inflammatory diseases, as large-scale data on many immune cell subsets can be readily generated from blood (Chen et al., 2016) and studied either in bulk (Aguet et al., 2020) or at single-cell resolution (Yazar et al., 2022). For instance, eQTL studies have played an important role in interpreting the results of genome-wide association studies (GWAS) of COVID-19 (Kousathanas et al., 2022). However, the majority of these studies are carried out in resting immune cells of healthy individuals, whereas the gene expression profile and gene regulatory landscape of immune cells is known to be altered dramatically when exposed to natural infection or artificial stimulation (Calderon et al., 2019; COMBAT Consortium, 2022; Fairfax et al., 2014), which may lead to important genetic mechanisms being missed in unstimulated cells.

Previous studies have investigated how eQTLs in immune tissue respond to stimuli (Cano-Gamez et al., 2020) and specifically pathogens in vivo such as influenza (Ye et al., 2020) and Salmonella (Alasoo et al., 2018), which highlighted new mechanisms of gene regulation by risk variants. There has been less work on organismal stimuli, such as a systemic infection. Large-scale studies in whole blood have shown that infectious disease status modifies the strength of a subset of eQTLs (Q. S. Wang et al., 2024), and targeted single-cell eQTL studies have demonstrated that the effect of certain COVID-19 risk variants on monocyte gene expression differs in COVID-19 patients (Edahiro et al. (2023) and that individual sepsis response may be driven by host genetic variation (Burnham et al., 2024). However, the role of infectious disease on risk-associated eQTLs across different infectious diseases and immune traits remains understudied.

In this study, we leverage new genotyping data from a previously described cohort, the COVID-19 Multi-omic Blood Atlas (COMBAT), to study how different infectious diseases and their severity modulate the impact of genetics on gene expression across a wide range of cell types. We combine genotype data with single-cell RNA-seq data from individuals defined as healthy and uninfected, as well as those unwell to varying degrees with COVID-19 and other causes of sepsis, who were consented and sampled as part of the COMBAT project. The aim of this study was to perform a genome-wide cis-eQTL association study and to discover new associations between inflammatory and infectious disease risk variants and gene expression in specific immune cells. We aimed to test whether infectious status modified the effect of risk variants on gene expression, and to shed new light on the mechanisms of gene regulation in infectious disease.

## Results

### Mapping cis-eQTLs across diverse cell types

We analysed single-cell expression of the transcriptome and genome-wide genetic data of 400,000 cells from 76 individuals of European ancestry from the COMBAT Consortium (COMBAT Consortium, 2022) (**Figure 1**). After quality control and imputation, we analysed 6,522,278 genetic variants.

**Figure 1:**
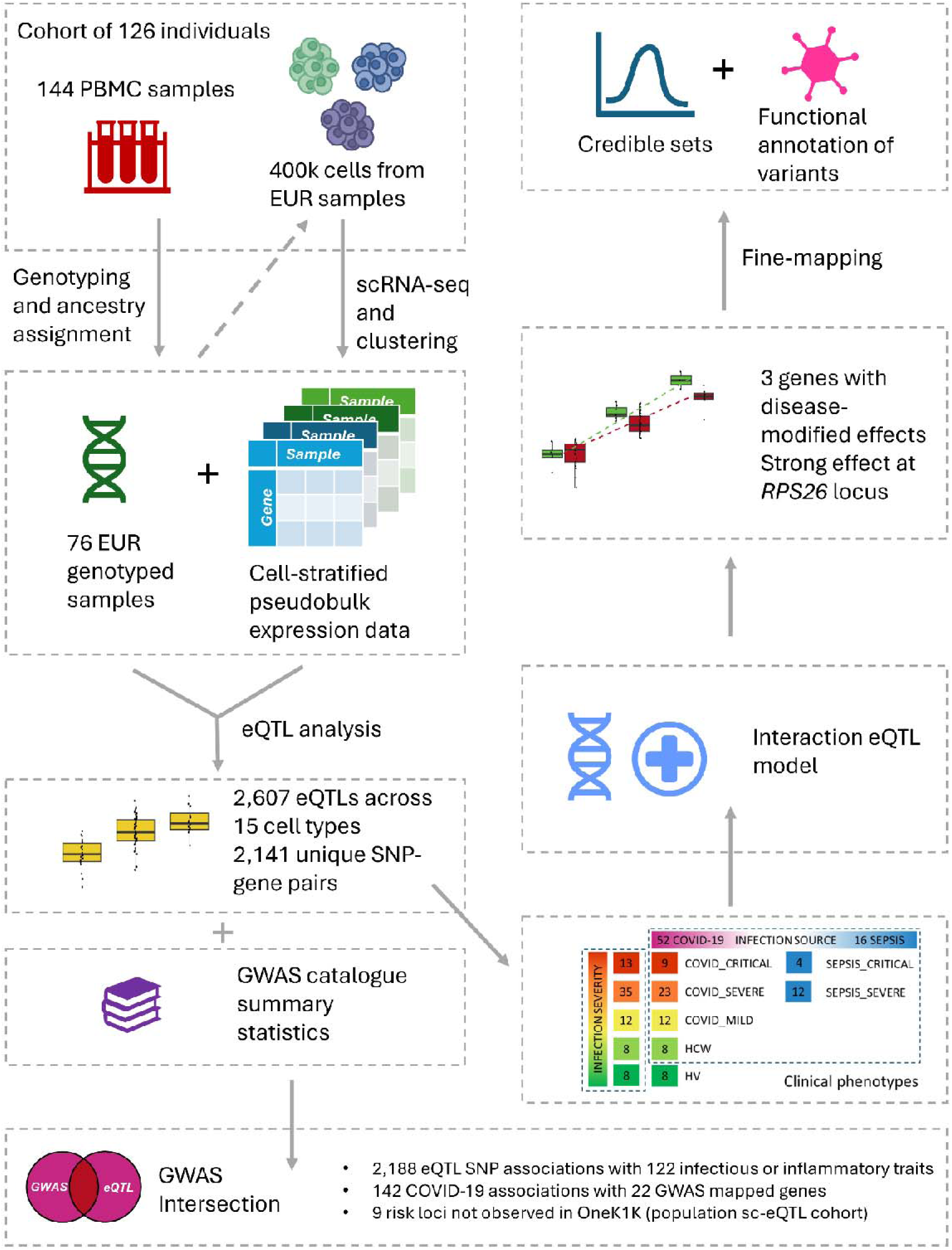
Schematic summarizing data, workflow and key results. scRNA-seq and clustering was previously described by the COMBAT consortium prior to this work (COMBAT Consortium, 2022). SNPs were genotyped and data were integrated for single-cell eQTL analysis

We tested *cis-*eQTL associations between covariate-corrected expression of genes expressed in more than 10% of samples and variants within 1Mb of the transcription start site (TSS) of each gene across 15 previously defined cell types (COMBAT Consortium, 2022) (**Figure 2a**). In total, we found 2,607 significant independent *cis*-eQTLs with 2,141 unique SNP-gene pairs (**Table S1**). We saw the most eQTLs in CD4+ T cells and classical monocytes. As expected, we found that the number of eQTLs discovered was positively correlated with the total number of cells assayed per cell type (**Figure 2b**). We observed that genes influenced by eQTLs (eGenes) are shared across cell types, with higher proportions of shared eGenes between functionally similar cell types (**Figures S1** and S**2**). For example, there are 250 eGenes found in both CD4+ and CD8+ T cells (27.7% of all the unique eGenes found in either of the two cell types are found in both) and 145 eGenes shared between CD8+ T and natural killer (NK) cells (34.4%). We noted a number of eGenes have previously been highlighted to play a potential role in COVID-19 infection (**Figure 2c**). These include an eQTL in the gene *NAPSA,* a candidate COVID-19 risk gene that plays a role in gas exchange in the lung (Degenhardt et al., 2022). In our data, we found that *NAPSA* also showed low but detectable expression in classical and non-classical monocytes (**Figure 2d**), where we found the eQTL signals (rs60182980, β_cMono_ = -1.20, se_cMono_ = 0.0932, p_cMono_ = 8.35×10^-19^, β_ncMono_ = -1.19, se_ncMono_ = 0.0792, p_ncMono_ = 9.64×10^-22^). Other candidate COVID-19 risk genes, including *ZGLP1* and *KANSL1* (Degenhardt et al., 2022; Kousathanas et al., 2022) also showed significant eQTLs in classical monocytes (*ZGLP1*, rs73510898, β = 1.44, se = 0.202, p = 1.56×10^-9^), NK cells (*KANSL1*, rs2696531, β = 1.00, se = 0.146, p = 4.51×10^-9^) and CD4+ T cells (*KANSL1*, rs17660167, β = 1.06, se = 0.104, p = 1.47×10^-14^ and rs62063286, β = 0.654, se = 0.105, p = 5.52×10^-8^). Expression of these genes in on Uniform Manifold Approximation and Projection (UMAP) plots is shown in **Figure S3**.

**Figure 2:**
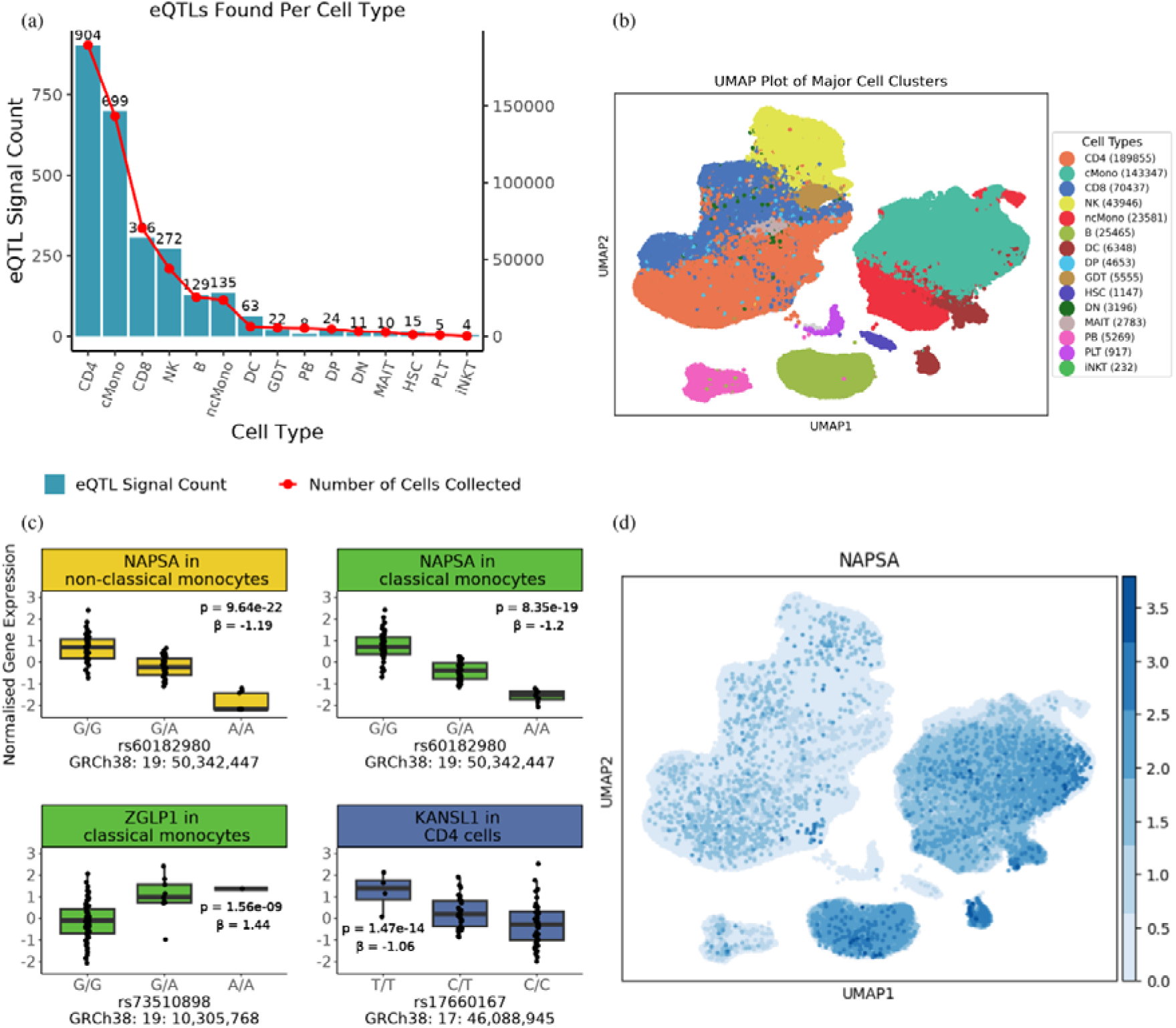
An overview of eQTL results. **a)** eQTL signal counts (y-axis left) and cell counts (y-axis right) across 15 cell types (x-axis) **b)** UMAP plot of 526,731 cells across 76 individuals with 15 annotated cell types. The abbreviations for each cell type are displayed in Table S8 with cell counts for each cell type **c)** eQTL boxplots for COVID-risk variants with risk-alleles on the right. The boxplots show the distribution of normalized gene expression (y-axis) across genotype categories (x-axis). The box represents the interquartile range (IQR), with the horizontal line indicating the median and the whiskers extending to 1.5×IQR. P-value and beta are derived from a linear model assessing the association between genotype and gene expression **d)** UMAP plot with differential expression of *NAPSA* (higher expression in B cells and monocytes)

### eQTL variants are enriched in many complex traits

Consistent with previous work, we found that the many of our eQTLs were in high linkage disequilibrium (R^2^ >0.8) with genome-wide significant loci associated with COVID-19, as well as infectious and inflammatory diseases including inflammatory bowel disease (Crohn’s disease and ulcerative colitis), rheumatoid arthritis, systemic lupus erythematosus and type 1 diabetes (**Figure 3a, Table S2**).

**Figure 3:**
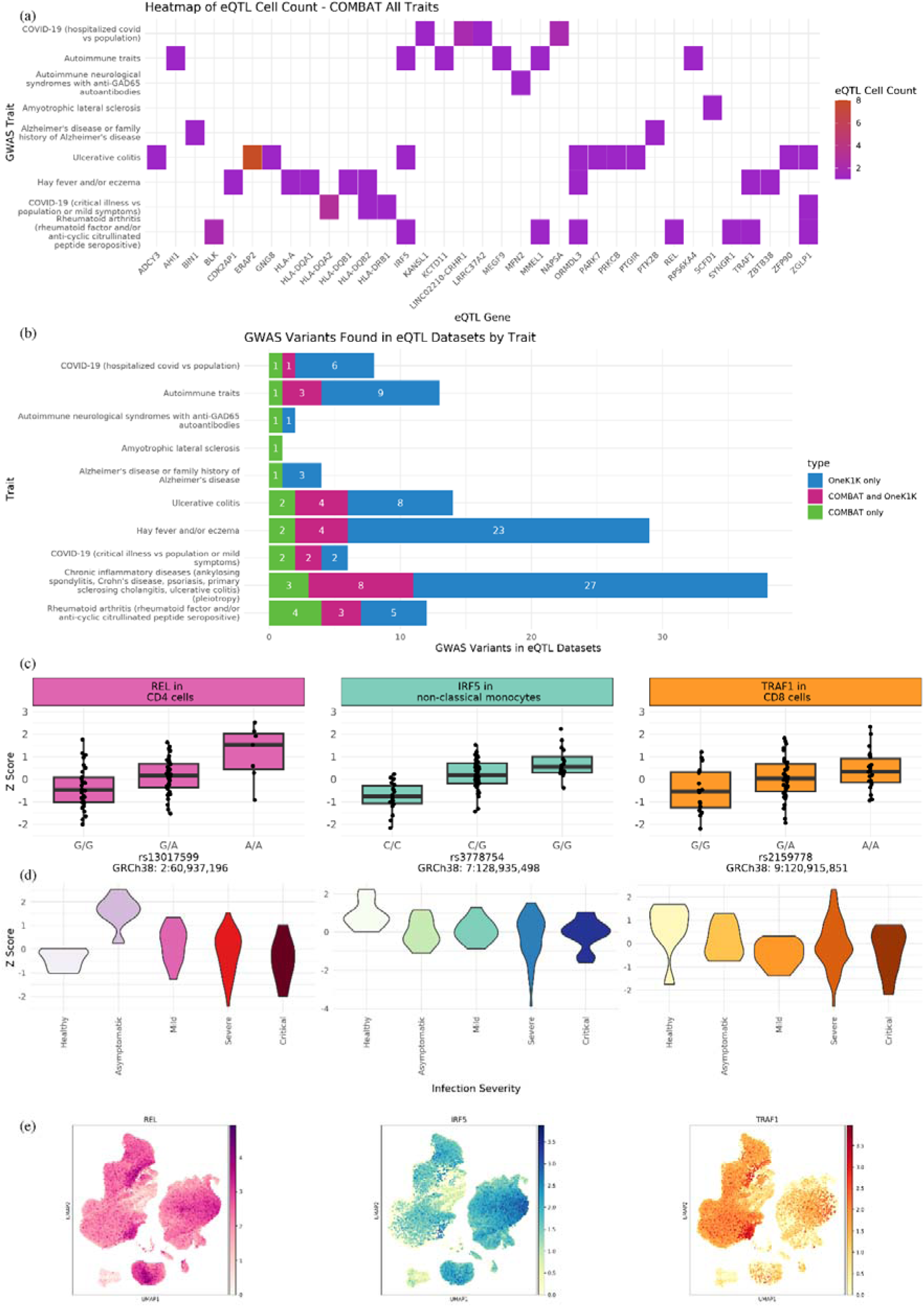
Colocalization of eQTL and GWAS variants. **a)** COMBAT eQTL SNPS colocalizing with GWAS risk variants for infectious and inflammatory traits according to the GWAS catalogue downloaded on 30/05/2024 (Cerezo et al., 2025) **b)** Overlap between lead SNPs of the eQTL analysis and GWAS traits (infectious and immune) in COMBAT and OneK1K **c)** Boxplots of strongest *REL*, *IRF5* and *TRAF1* eQTLs. The boxplots show the distribution of normalized gene expression (y-axis) across genotype categories (x-axis). The box represents the interquartile range (IQR), with the horizontal line indicating the median and the whiskers extending to 1.5×IQR **d)** Violin plots showing expression of *REL*, *IRF5* and *TRAF1* stratified by infection severity, over the categories healthy (healthy volunteers), asymptomatic (individuals with no symptoms but a positive COVID-19 test), Mild (hospitalized with mild COVID-19 or sepsis infection), Severe (hospitalized with severe COVID-19 or sepsis infection and requiring no ventilation or non-invasive ventilation) and Critical (hospitalization with critical COVID-19 or sepsis infection requiring intubation) **e)** Single-cell expression of *REL*, *IRF5*, *TRAF1* reflected on a UMAP plot

It is likely that many of these eQTLs are not related to infection and would also be found in healthy individuals. To assess this, we compared our results to those from OneK1K (**Table S3**), a large, publicly available genotyped cohort that was not ascertained for disease cases (Yazar et al., 2022). As expected, we found a large overlap between the eQTLs in infectious disease cases and unascertained samples; 1,115/1,513 (73.7%) of our eGenes were also eGenes in OneK1K. We found that the majority of our trait-associated eQTLs were also reported in OneK1K (345/430, 80.2%).

Interestingly, our infection dataset also uncovered trait-associated eQTLs that were not present in the general population dataset (**Figure 3b**, **Figures S4-S6**). Here we highlight three genes: *REL* in CD4+ T cells, *IRF5* in non-classical monocytes and *TRAF1* in CD8+ T cells. These risk genes showed association with GWAS risk variants in the infectious disease cohort but not in the general population cohort, despite being measured in immune cell subsets in both studies (**Figure 3c**). These are all established risk genes for inflammatory diseases with Open Targets (Ghoussaini et al., 2021; Mountjoy et al., 2021) L2G scores > 0.6: *REL* is a myeloid checkpoint gene associated with rheumatoid arthritis, *IRF5* play a role in interferon-mediated response to viral infection and is associated with rheumatoid arthritis and inflammatory bowel disease, and *TRAF1* mediated TNF-receptor-family signalling and is associated with rheumatoid arthritis. Notably, all three genes were differentially regulated by infection severity; *IRF5* and *TRAF1* decrease as severity increases, while *REL* is initially upregulated with infection and subsequently decreases (**Figure 3d**) and are differentially expressed in immune cell subsets of major groups, showing the importance of working at single-cell resolution (**Figure 3e**). These genes have all been previously shown to be affected by genetic variation in whole blood in the general population (Aguet et al., 2020), but our dataset suggests they may play a role in specific immune cell subsets in infectious disease.

### Modification of eQTL signals by disease status across multiple cell types

We next tested whether any of the 2,607 eQTL effects showed evidence of interaction with infection status, i.e. whether the genetic variants have a varying effect on gene expression depending on the disease severity or source of infection. For infection source, we tested the effect of hospitalization for COVID-19 vs. for all-cause sepsis (excluding healthy controls). For infection severity, we defined a linear scale from 0 to 4 (0: heathy, 1: asymptomatic, 2: hospitalization with mild infection, 3: hospitalization with severe infection and requiring no ventilation or non-invasive ventilation, 4: hospitalization with critical infection requiring intubation, see Figure 1). In total, we identified three genes where the effect size was modified by infection status (source or severity), with effect modification across several key cell types involved in immune response (Table 1). One of the genes was in the MHC region of chromosome 6 (HLA-DQB2), which was modified by infection severity, and two were outside the MHC (RPS26 and ADAM10) impacted by a mixture of source and severity across cell types (Figure 4).

**Figure 4:**
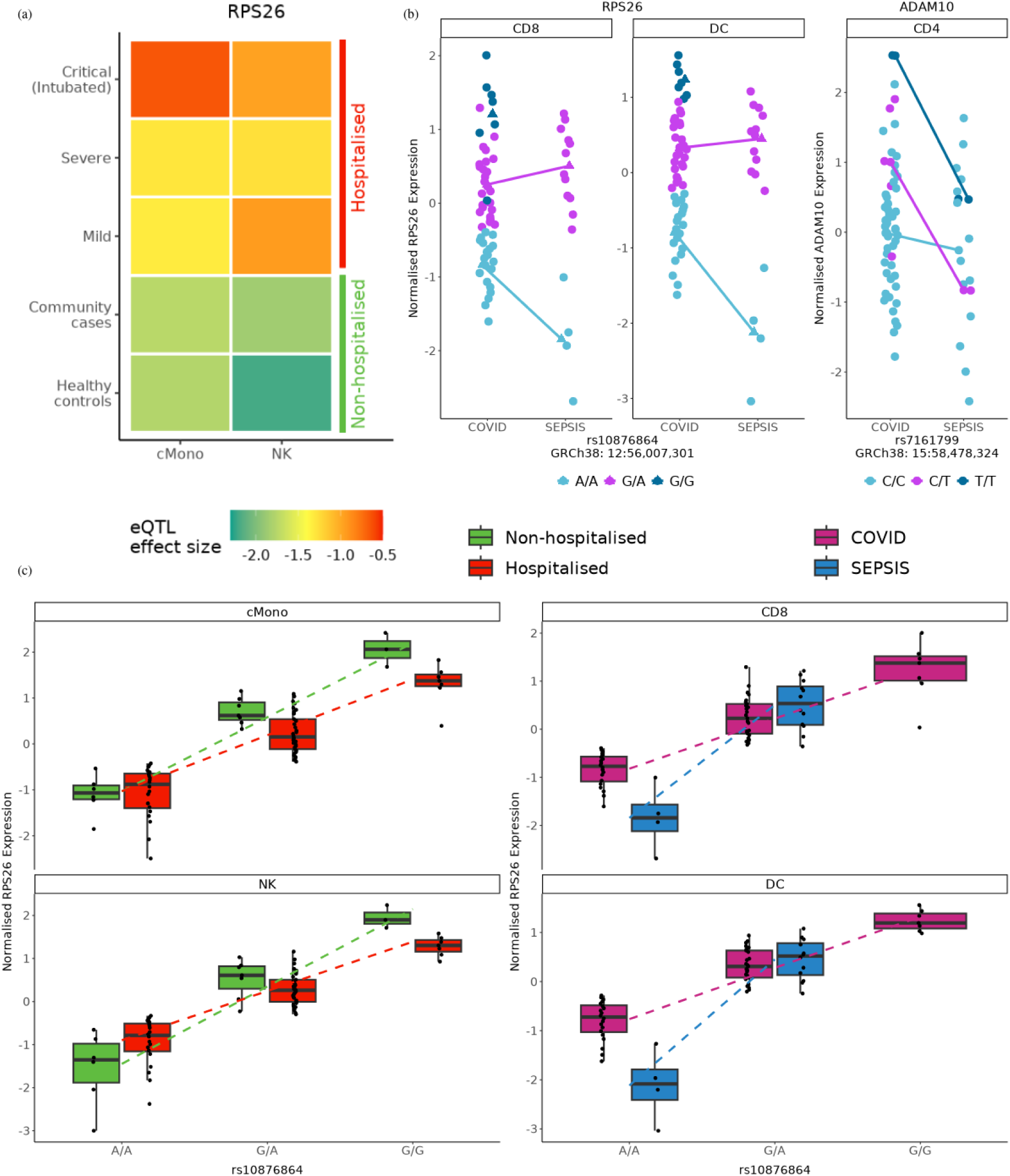
Context-modified eQTLs *RPS26* and *ADAM10*. **a)** Heatmap shows the magnitude of effect size of rs10876864-*RPS26* decreases as infection severity increases in classical monocytes and NK cells. Heatmap colours denote betas of eQTL effect from the regression model **b)** Plot shows source of infection modifies eQTL effect at *RPS26* (CD8+ and DC) and *ADAM10* (CD4+) loci. Normalized gene expression was stratified by two phenotypes (COVID-19 and Sepsis), coloured by individuals’ genotype of the lead eQTL variant for *RPS26* and *ADAM10*. **c)** Genotype (x-axis) association with normalized *RPS26* expression (y-axis) modified by severity and source of infection. The box represents the interquartile range (IQR), with the horizontal line indicating the median and the whiskers extending to 1.5×IQR. The two boxplots on the left show the eQTL effect in green for non-hospitalized (healthy and asymptomatic) and red for hospitalized (mild, critical and severe infection) in cMono and NK cells. The two boxplots on the right show the eQTL effect in pink for COVID-19 and blue for sepsis for CD8+ and dendritic cells (DC). A/A genotypes show increased expression in hospitalized individuals whereas *RPS26* expression decreases in hospitalized G/G and heterozygous individuals. A similar relationship is seen with COVID-19 vs sepsis infection.

**Table 1:** A summary of the disease-specific eQTL results. Cell type, chromosome, gene, associated variant with effect allele and GRCh38 position, phenotype comparison model, interaction beta value, false detection rate (eQTL p-values were corrected using Benjamini-Hochberg correction).

| Cell | Chr | Gene Name | SNP | Infection Status | Interaction Beta | Adjusted p-value |
| --- | --- | --- | --- | --- | --- | --- |
| cMono | 6 | *HLA-DQB2* | rs3104367-T<br>(6: 32,635,710) | Severity | -0.450 | 0.004 |
| NK | 12 | *RPS26* | rs10876864-G<br>(12:56,007,301) | Severity | 0.264 | 0.028 |
| cMono |  |  |  |  | 0.187 | 0.027 |
| CD8 |  |  |  | Source | 1.304 | 0.027 |
| DC |  |  |  | Source | 1.340 | 0.003 |
| CD4 | 15 | *ADAM10* | rs7161799-C<br>(15:58,478,324) | Source | 1.168 | 0.049 |

The strongest infection-modified eQTL effect was at the *RPS26* locus, a gene which is widely expressed across cell types (**Figure S4**) and is strongly regulated by variant rs10876864. The effect size of the eQTL decreases with increasingly severe infection, with a notable change between non-hospitalized and hospitalized individuals in classical monocytes (cMono) and NK cells (**Figures 4a and 4c**).We also observed a larger effect size in sepsis than COVID-19 in CD8+ T and dendritic cells (**Figures 4b and 4c**).

We next used statistical fine-mapping to identify candidate causal variants for the eQTLs in *RPS26* and *ADAM10* (we did not fine-map the HLA eQTL due to extensive linkage disequilibrium in the region) and used this to investigate potential regulatory effects of these variants (**Table S4**). We were able to fine-map the *RPS26* eQTL to two candidate variants (rs1131017 and rs10876864) in high linkage disequilibrium (R^2^ = 0.992 in Europeans), both with a posterior probability of being causal of 50%. We fine-mapped the *ADAM10* eQTL to a 95% credible set of three variants, of which one (rs7161799) had a substantially larger posterior probability (93%).

We annotated all variants within the credible sets after fine-mapping with information on chromatin state, transcription factor motifs and functional SNP scores (**Table S5**). The two lead fine-mapped *RPS26* variants rs1131017 and rs10876864 are reported by RegulomeDB to be in open chromatin regions in the cell types where the eQTL was detected, with one (rs1131017) in an active transcription start site in all cell types. The top *ADAM10* eQTL candidate causal variant was predicted to disrupt the binding motifs of multiple transcription factors.

## Discussion

We report genome-wide eQTL across 15 key immune cell types with and without context-modified effects, with particularly large effects in CD4+ T cells and classical monocytes. Consistent with previous observations from blood cells in healthy individuals (Yazar et al., 2022), we observed effects of COVID-19 risk variants on expression of multiple genes across multiple cell types of infectious disease patients. We also found a number of genes implicated by both our eQTL results and published GWAS (**Table S2**) that were not observed in a population cohort including *REL*, *IRF5*, and *TRAF1*, all of which are differentially regulated with infection severity in our results. This may imply differential gene regulation between infected and healthy cohorts for example via disease-specific regulatory elements and shows that disease-specific single-cell datasets can show new roles for established eQTLs.

There were few context-specific eQTLs; most eQTLs are not detectably modified by infection. The eQTL map of a COVID-19 patient is hard to distinguish from the eQTL map of a healthy person at this sample size. One can find context-modified eQTLs, but they are not context-specific i.e. they are not simply present or absent depending on infection status. Rather, infection can amplify or reduce the impact of a genetic variant on gene expression.

We do not see any strong intersections between COVID-19 genetics and context-modified eQTL hits. However, the context-modified eGenes we found did generally have known roles in the pathogenesis of COVID-19. The *ADAM10* gene is expressed in many tissues including the central nervous system, immune cells, and respiratory tract; it is broadly expressed in lung tissue and is associated with promoting SARS-CoV-2 cell entry (Jocher et al., 2022). Although not universally and highly expressed across cell types in our data (**Figure S3**), implying some interaction effects could be missed, the results reflect the biology and known associations of these genes with COVID-19.

The strongest eQTLs, both with and without infection interaction, were found near the *RPS26* locus (variant rs10876864) which showed convincing context-modified effects in four cell types including CD4+ T cells and classical monocytes, with reduced eQTL effects sizes in hospitalized patients. We fine-mapped this association to two candidate causal variants (rs10876864 and rs1131017), both of which were in active regulatory regions in immune cells. This is a known risk locus for type 1 diabetes (Forgetta et al., 2020, Burton et al., 2007; Márquez et al., 2018) and rheumatoid arthritis (Laufer et al., 2019; Okada et al., 2013). The gene *RPS26* is known to play a role in COVID-19, as a marker of disease severity and stage in T cells (Ren et al., 2023), (Vázquez-Jiménez et al., 2021). The risk variant rs10876864 has not previously been reported as a genome-wide significant locus for COVID-19, but it has a marginally significant association with hospitalized COVID19 (HGI analysis B2, release 7, p = 2.81 × 10^-3^, The COVID-19 Host Genetics Initiative (2020)).

Our results for *RPS26* and *ADAM10* implicate specific fine-mapped genetic associations that are both associated with human disease and lie in activate regulatory elements that are impacted by infection in specific cell types. They thus provide good candidates for future mechanistic experiments for studying gene regulation in infectious disease using techniques such as CRISPR base editing (Schmidt et al., 2024).

This study has several important limitations. All samples were derived from whole blood rather than disease-relevant tissues, which may have obscured tissue-specific regulatory effects. The study also has limited statistical power due to a relatively small sample size (n = 76). The cohort was somewhat imbalanced, with relatively few sepsis patients and healthy controls compared to the number of COVID-19 cases. Finally, although immune responses and gene regulatory effects likely evolve throughout infection and recovery, the study was restricted to cross-sectional sampling and lacked sufficient longitudinal data to capture dynamic regulatory changes over time. These limitations should be addressed in future studies with larger, more balanced, and longitudinally sampled cohorts.

In conclusion, we have identified several immune cell sub-populations including CD4+ and CD8+ T cells and classical monocytes, in which eQTL effects are present both with and without an interaction with infection. We have learned that, for a small number of variants, the effect of genotype on expression differs depending on infection status and disease severity. However, the magnitude of effects is relatively small compared to effect of genotype on expression in general and the effect should be considered a modification rather than specific to the presence or absence of disease. Some genetic variants modified by disease include those that also play a role in inflammatory disease including type 1 diabetes and rheumatoid arthritis. These results emphasise the genetic contribution to individual immune responses and demonstrate the importance of expanding these studies to further understand host defence mechanisms in the context of infectious disease.

## Methods

### Sample collection and genotyping

For COVID-19 patients (hospitalized patients and healthcare workers) and acute sepsis patients, whole blood was collected in Tempus blood RNA tubes, and DNA and RNA were extracted using firstly a Total RNA Purification kit (Norgen), followed by a Monarch Genomic DNA Purification Kit (New England Biolabs) used on the first supernatant from the RNA extraction step. Manufacturer protocols were followed for both kits. For healthy volunteers and non-acute sepsis patients, DNA was extracted from buffy coat samples. 144 samples were collected for genotyping (**Table S6**). Samples were genotyped on two plates (target of 500ng per well, achieved range 260-500ng), using the Illumina Infinium Global Screening Array (version 3). Genotypes were called using GenomeStudio v2.0.4.

### Genotype quality control, ancestry assignment and imputation

A total of 144 individuals were genotyped for 730,059 variants. For quality control (QC), we removed samples based on sex mismatches, low call rate (<98%) and heterozygosity outliers. We removed SNPs based on low call rate (<98%), low minor allele frequency (0.02) and Hardy-Weinberg equilibrium p-value (p<0.001). We further removed insertion/deletion variants and copy number variations (m=1,089) and strand-ambiguous SNPs (m=5,689).

We next carried out a principal component analysis (PCA) to assign continental ancestry to our samples. We intersected our samples with 1000 Genomes genotypes (phase 3, v5a.20130502, (Auton et al., 2015) and carried out linkage disequilibrium thinning (using Plink settings of window size=50, shift=5, VIF=0.2, resulting in 202,954 variants). PC axes were generated using 1000 Genomes samples, with COMBAT individuals projected onto these axes (**Figure S7**). Each individual was assigned to one of the five major continental ancestry groups from the 1000 Genomes (African/AFR, American/AMR, East Asian/EAS, European/EUR, South Asian/SAS) by calculating the sum of absolute differences between each COMBAT individual and each of the five populations across the first 10 PCs (after normalizing each PC to have variance 1 across 1000 Genomes individuals), and assigning each COMBAT individual to its nearest 1000 Genomes population on this measure. Each of the major populations were manually inspected to identify outliers. In total, we identified 115 European, 10 East Asian, 6 African and 5 South Asian samples, plus 4 outlier samples.

The heterozygosity and Hardy-Weinberg tests were only carried out in European individuals. Samples were also checked for duplicated or closely related samples (Identity by descent proportion < 0.2), and for consistency with called genotypes called from previously published bulk RNA-seq (COMBAT Consortium, 2022). After QC, a total of 140 individuals and 456,247 variants remained for imputation. All QC was carried out in Plink v1.90 (Chang et al., 2015).

Imputation was carried out on all QC-passing samples using the TOPMed reference set via the TOPMed Imputation Server (Taliun et al., 2021) and using the 1000 Genomes reference set via the Michigan Imputation Server (Das et al., 2016). Reference alleles and strand were assigned, and data formatted for imputation, using HRC-1000G-check-bim v4.30 (https://www.well.ox.ac.uk/~wrayner/tools/). Data was phased with Eagle2 (Loh et al., 2016) and imputed using minimac4 (Howie et al., 2012). Output was filtered to remove variants with predicted R^2^ < 0.7 and minor allele frequency < 0.05, and RSIDs were assigned from dbSNP (build 151), both using bcftools (Danecek et al., 2021) (v1.10.2). Imputation quality was checked using the default leave-one-out metrics produced by minimac4, showing higher performance for the TOPMed reference set even at low allele frequencies compared to the 1000 Genomes reference panel (**Figure S8**), and so for all analyses we used this imputed dataset with 6,522,278 SNPs.

### Pseudobulk data manipulation: processing and filtering

We used the quantified scRNA-seq count data described in the previous COMBAT publication (COMBAT Consortium, 2022). We used 15 of the previously described “major subset” cell-type annotations (**Table S8**), excluding RET and mast cells due to low cell numbers, and produced summed pseudobulk counts for each sample for each cell type. Individuals who were not genotyped and those who clustered with non-European reference populations from the 1000 Genomes project were excluded. For individuals with samples taken at multiple time points, the first sample was used. Pseudobulk expression values were converted to counts per million (CPM) then quantile normalized and scaled to mean 0 and variance 1 across genes. Genes with non-zero CPM in <10% of samples or an average CPM across samples of <1 were removed, with between 4,482 (PLT) and 10,596 (CD4) genes retained across the remaining cell types.

We calculated principal components of gene expression for each cell type, using the top 3000 most variable genes, which were used as covariates for eQTL mapping.

### eQTL mapping

We used tensorQTL (Taylor-Weiner et al., 2019) to map eQTLs across all variants in the imputed genotype data and all genes passing quality control to test for eQTLs in each cell type. We used the *cis* mode to identify eGenes with significant (FDR < 0.05) associations with at least one variant, which uses permutation ro produce multi-testing-corrected p-values per gene followed by the Benjamini-Hochberg procedure to calculate adjusted q-values per gene. We used *cis-nominal* mode with a minor allele frequency cut-off off 0.05 to obtain summary statistics for each variant-gene pair (for all variants within 1Mb of the transcription start site of the gene). Finally, we ran *cis-independent* mode which uses the *cis* results to find genes with at least one secondary (conditionally independent) cis-QTL. The model used by tensorQTL (Equation 1) describes eQTL signals in each cell type, where there is no interaction between genotype and disease.

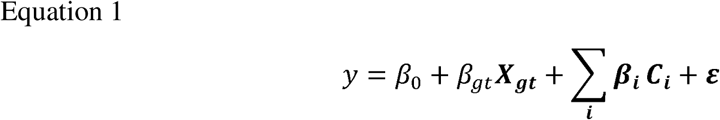

where the sum of covariate terms ***c*** is over

*i* ∈ {*age, sex, disase,* 2 *genotype PCs,* 5 *expression PCs*}.

Five expression PCs and two genotype PCs were chosen based on visual inspection of cumulative variance explained plots.

### GWAS colocalization

We tested whether any of our conditionally independent eQTL variants were in, or were in high linkage disequilibrium (LD, R^2^ > 0.8) with variants in, the GWAS catalogue (downloaded 30/05/2024) (Cerezo et al., 2025). LD was calculated in the 1000 Genomes European samples using plink1.9.

We categorized results as either inflammatory or infectious traits and annotated all entries with in-phase haplotypes, GWAS and eQTL summary statistics and apparent associations (**Table S2**). When the same trait was represented in multiple studies in the GWAS catalogue, we kept only the study with the largest sample size. For comparison, we also repeated this process with the OneK1K dataset (Yazar et al., 2022) to compare our results to a large population cohort not selected for infectious disease status (**Table S3**).

We used available resources including the GTEx Portal (accessed 21/10/2024, Aguet et al. (2020)) and the Open Targets Genetics (Ghoussaini et al., 2021; Mountjoy et al., 2021) to investigate the function and relevance of these genes.

### Disease-specific eQTL

To test for disease-specific eQTL, we categorized individuals based on their clinical phenotype at the time of sample collection. Each individual was assigned a numerical ‘infection score’ (0 for healthy control, 1 for infected but asymptomatic healthcare workers, and 2-4 for mild, severe, and critically ill hospitalized cases). We further categorized these into ‘non-hospitalized’ (0-1) and ‘hospitalized’ (2-4). We also tested infection source, categorising individuals as COVID-19 or sepsis (excluding healthy controls). See **Figure S9** for an illustration of this schema.

We tested for interaction between genotype and these infection status measures using a linear model implemented in R (R version 4.3.2). The null model H_0_ is described by Equation 1 (with no interaction term). The alternative model H_1_ (Equation 2) describes an eQTL at which there is an interaction between genotype and disease:

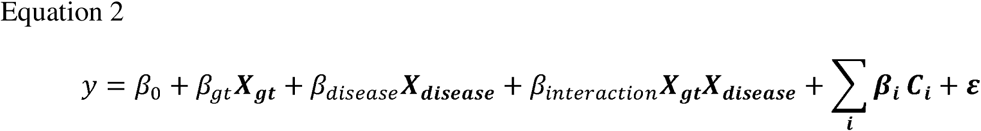

where the sum of covariate terms ***c*** is over

*i* ∈ {*age, sex,* 2 *genotype PCs,* 5 *expression PCs*}.

For each of the cis-independent variant-gene-cell combinations in which a signal was found by tensorQTL, we ran an ANOVA model to compare H_0_ and H_1_ and calculate summary statistics for each combination of variant, gene, cell type and phenotype categorization (Equation 3).

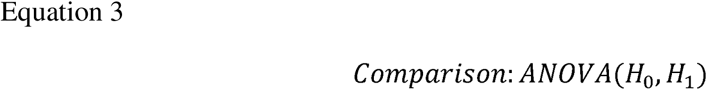

The p-values generated by ANOVA were adjusted using the Benjamini-Hochberg procedure with a threshold of FDR < 0.05.

### Fine-mapping of interaction eQTL variants

We performed fine-mapping of each eQTL variant with a context-specific effect (not including those regulating expression of HLA genes). We used the *cis-nominal* mode within tensorQTL to obtain nominal summary statistics for each cell type in which there was a disease-genotype interaction effect. We used the approached described by Maller et al. (2012) to fine-map variants in LD with the lead eQTL variant using a threshold of R^2^>0.4 according to the LDproxy (accessed 06/03/2024) function of LDlink (Machiela & Chanock, 2015). We performed a sensitivity analysis to compare different values of the prior variance on the effect size (**Figure S10**, **Table S7**), and selected a prior variance of 20% of variance in expression in accordance with susieR (G. Wang et al., 2020). We calculated posterior probabilities of causality for each variant and defined credible sets as the smallest set of variants for which the posterior inclusion probabilities summed to greater than or equal to 0.95.

We used RegulomeDB (Boyle et al., 2012) and FORGEdb (Breeze et al., 2024) to annotate all variants within the credible sets with information on chromatin state, transcription factor motifs and functional SNP scores. Chromatin states were investigated both for PBMCs and for the cell type in which the context-specific eQTL was reported. We used the chromatin state categories described by Ernst & Kellis (2017) to identify variants with potential regulatory effects.

## Data and code availability

CITESeq single-cell expression data containing cell-, gene- and individual-level metadata (COMBAT-CITESeq-DATA.h5ad) are publicly available at: https://zenodo.org/record/5139561/

Other patient data are available under controlled access at the European Genome-phenome Archive (EGA) under the study ID EGAS00001005493. Genotype calls and and imputed genotype data will be made available via this EGA study ID on publication.

Data and code to reproduce figures are available at: https://github.com/astleyjennie/COMBAT_eQTL

## Author contributions

Conceptualization: JA, JK, AM, YL, LJD; Data curation: JA, AK, BH, CD, SS; Formal analysis: JA, BH, CD, SS; Funding acquisition: JK, AM, YL, LJD; Investigation: AK, AM; Methodology: JA, YL, LJD; Project administration: JK, YL, LJD; Supervision: SS, JK, AM, YL, LJD; Writing – original draft: JA, BH, YL, LJD; all authors contributed to the review and editing of the manuscript.

## Acknowledgments

Research supported by the University of Oxford COVID-19 Research Response Fund and NIHR Oxford Biomedical Research Centre. JA is supported by MRC-studentship award MR/N013468/1(project 2748611). YL is supported by a Kennedy Trust KTRR Senior Research Fellowship (KENN202109). LJD and SS are supported by the Kennedy Trust for Rheumatology (KENN71803). LJD and BH are supported by the Wellcome Trust (208750/Z/17/Z).

