## Supplemental Figures for "Single-cell eQTL mapping identifies disease-specific genetic control of COVID-19 infection"

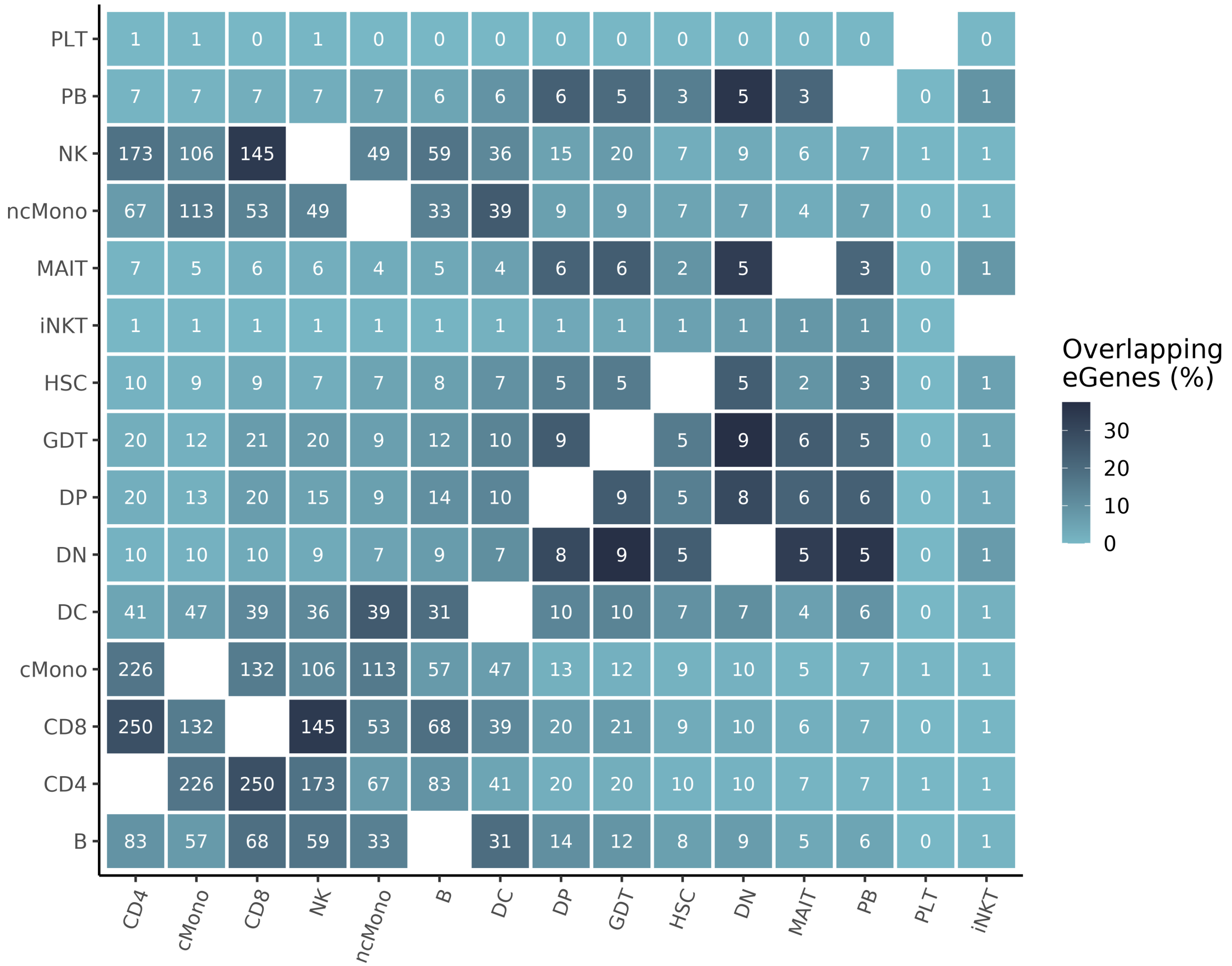


**Supplementary Figure 1:** Heatmap showing overlapping eGenes for each cell type. Colour: Jaccard index percentage (100  size of intersection over size of union), label: raw number of eGenes.

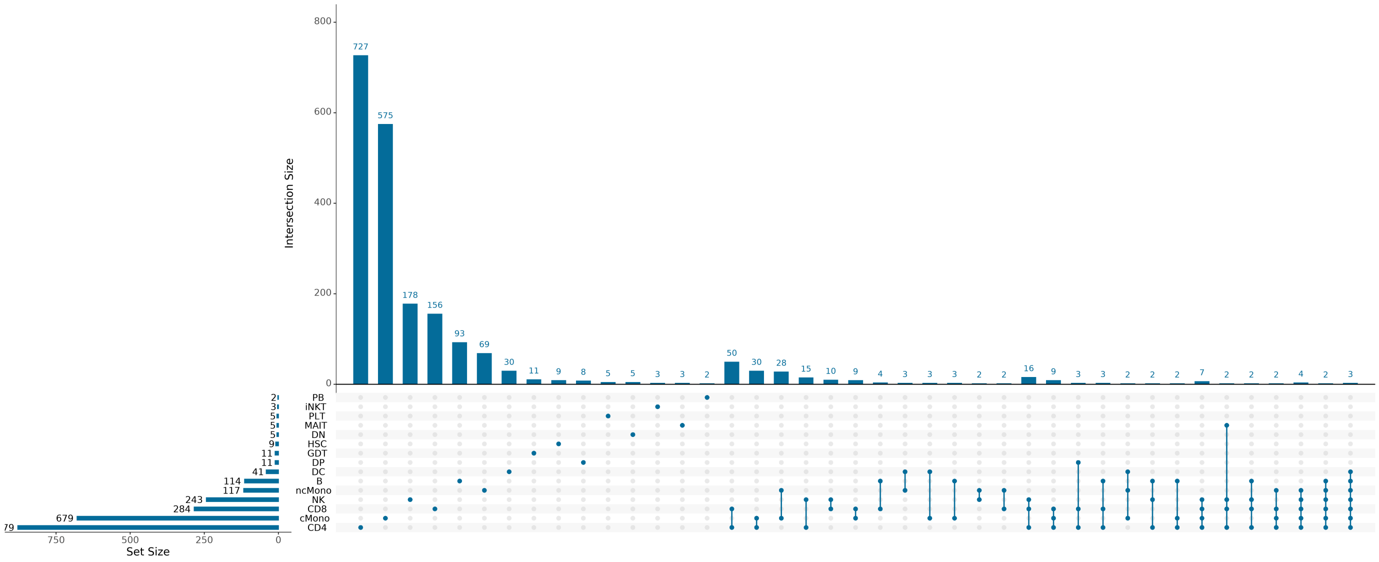


**Supplementary Figure 2:** UpSet plot showing all overlapping eGene results between cell types. Horizontal bars show how many independent Gene−SNP pairs are in each individual cell type. Vertical bars show how many Gene−SNP pairs are shared across the cell types indicated by the connected dots below each bar. Vertical bars of only one Gene−SNP pair have been removed for visual clarity.

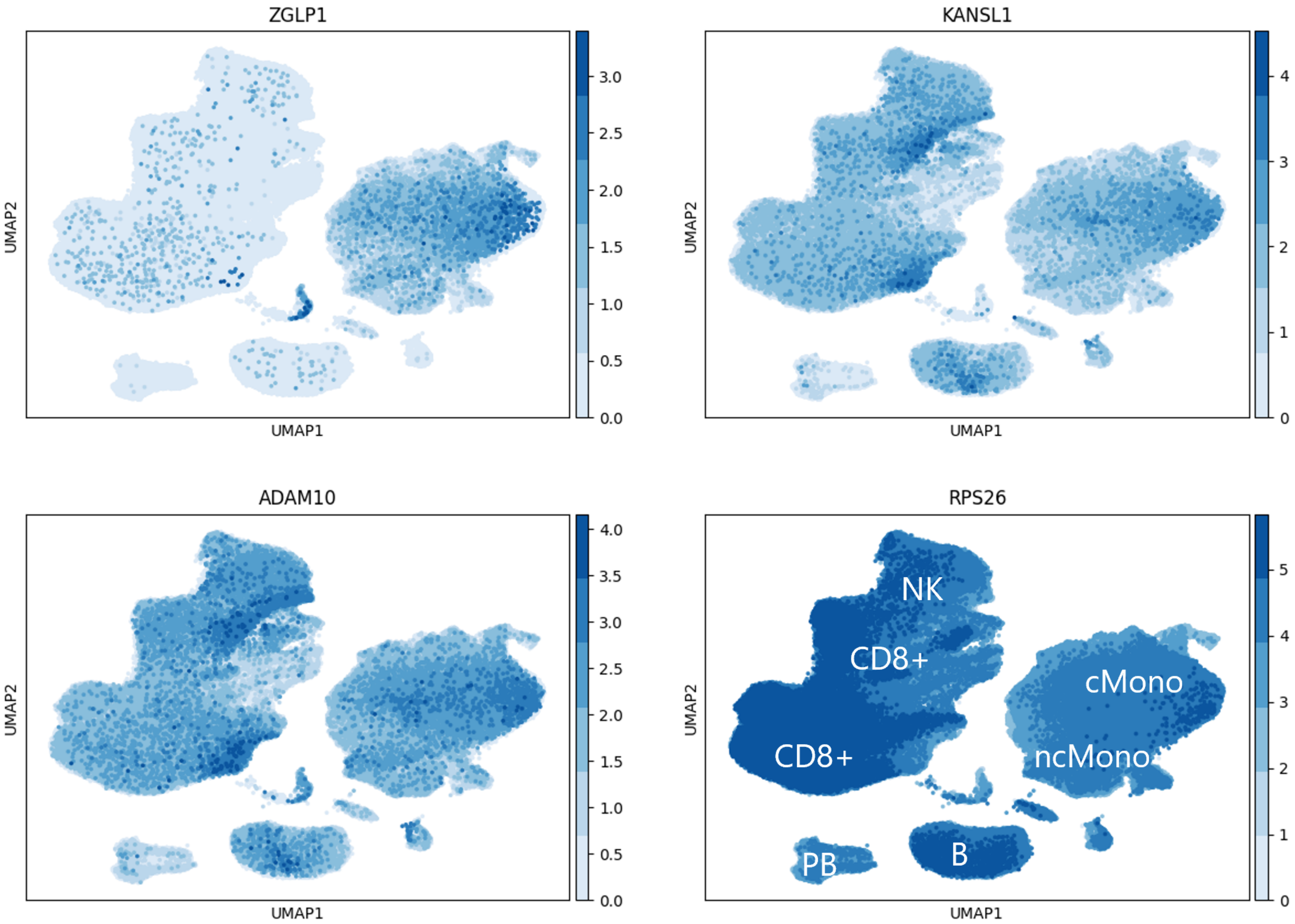


**Supplementary Figure 3:** UMAP plot with differential expression of *ZGLP1*, *KANSL1*, *ADAM10*, *RPS26* . Main cell types are shown on bottom right plot. UMAP of all cell types shown in Figure 2.

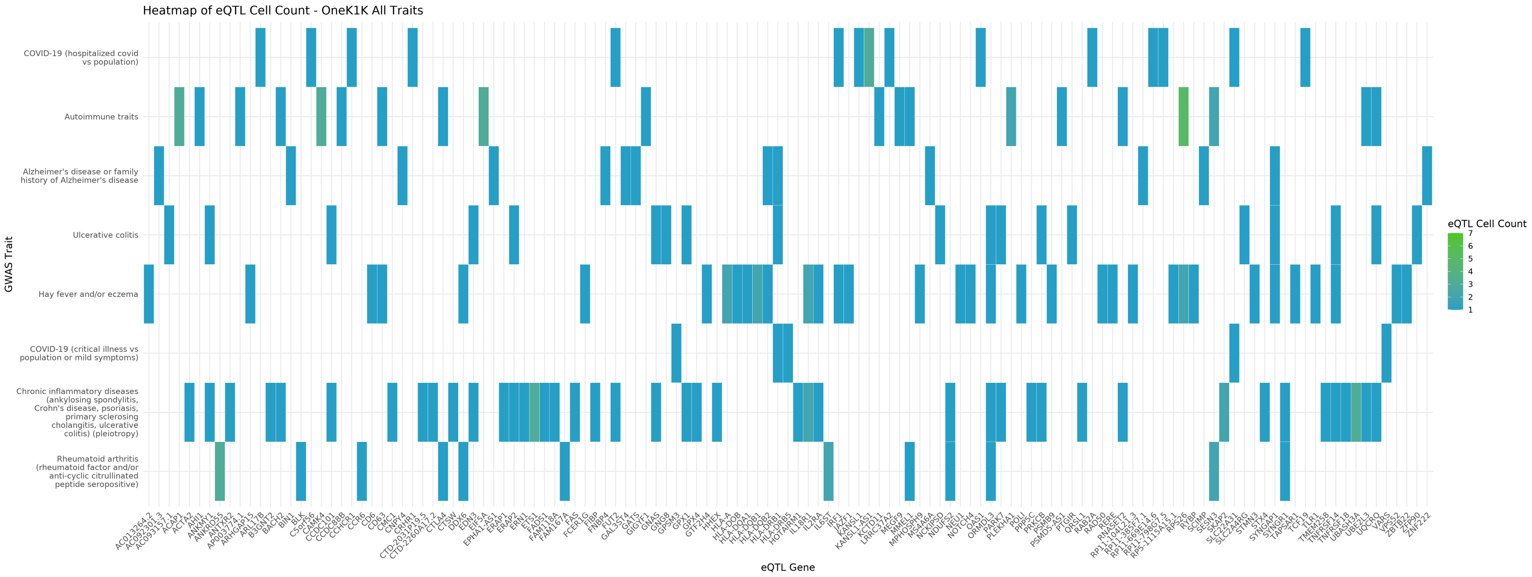


**Supplementary Figure 4:** OneK1K eQTL SNPs colocalising with GWAS risk variants for infectious and inflammatory traits. The x−axis shows the eGene associated with the eQTL variant and the y−axis shows the trait associated with the variant (or a variant in high LD, R2 > 0.8) in GWAS. The colour of the tile shows the number of cell types in which the eQTL was found.

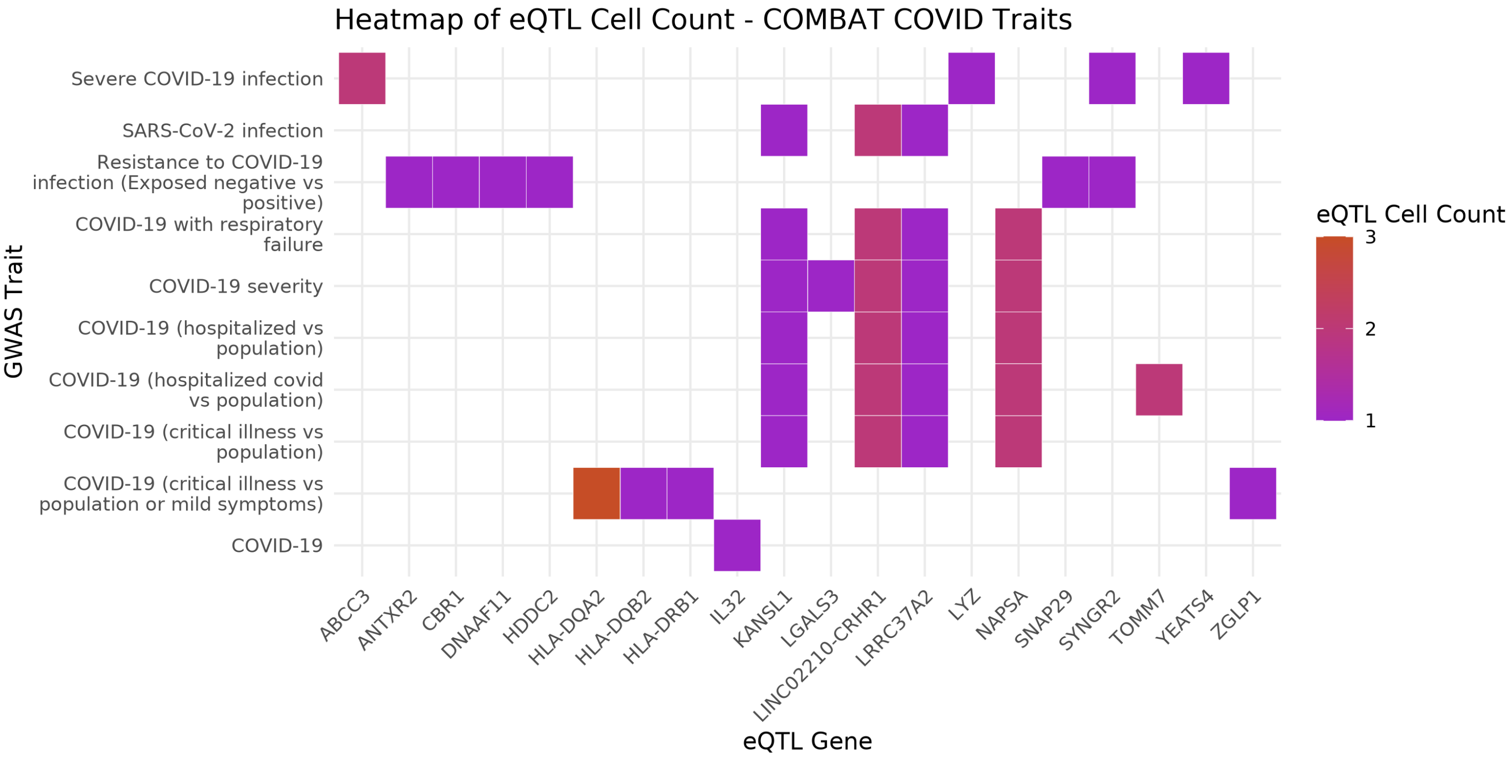


**Supplementary Figure 5:** COMBAT eQTL SNPs colocalising with GWAS risk variants for COVID−19 traits (severity, hospitalisaton and resistance of disease as well as SARS−CoV−2 infection). The x−axis shows the eGene associated with the eQTL variant and the y−axis shows the trait associated with the variant (or a varian in high LD, R2 > 0.8) in GWAS. The colour of the tile shows the number of cell types in which the eQTL was found.

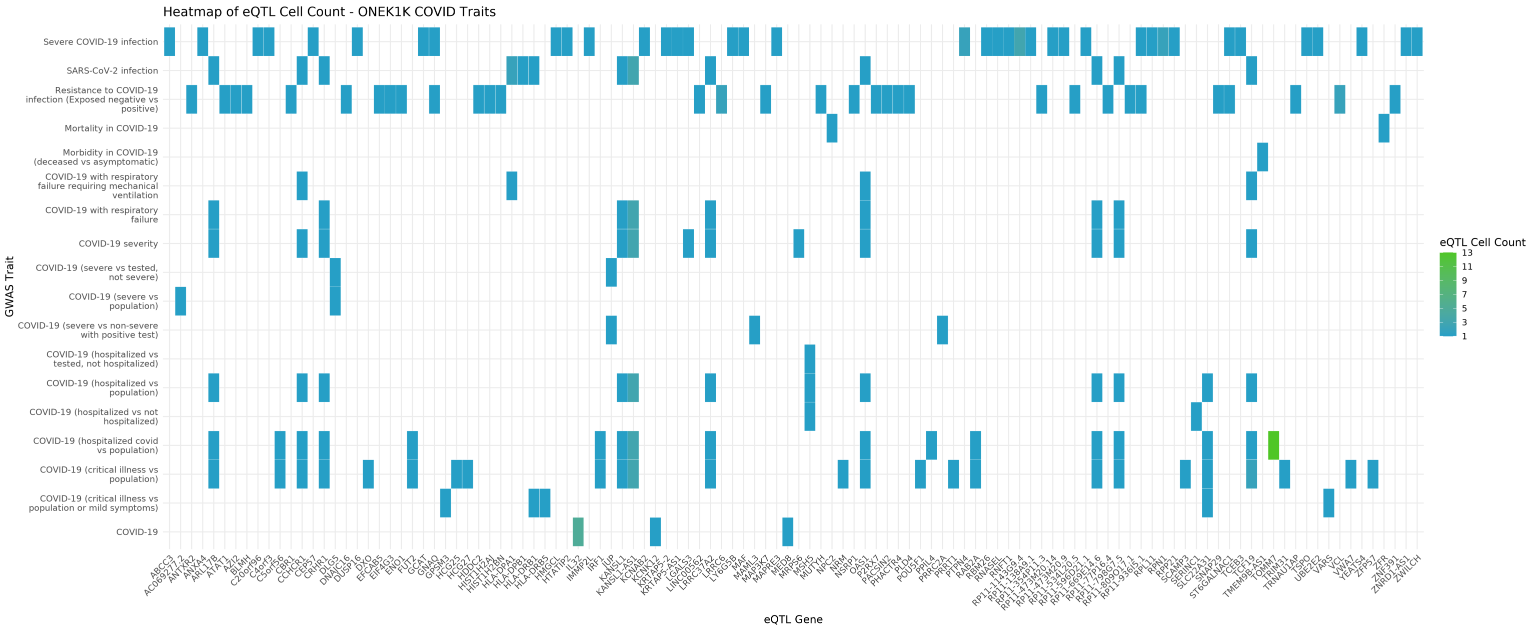


**Supplementary Figure 6:** OneK1K eQTL SNPs colocalising with GWAS risk variants for COVID−19 traits (severity, hospitalisaton and resistance of disease as well as SARS−CoV−2 infection). The x−axis shows the eGene associated with the eQTL variant and the y−axis shows the trait associated with the variant (or a variant in high LD, R2 > 0.8) in GWAS. The colour of the tile shows the number of cell types in which the eQTL was found.

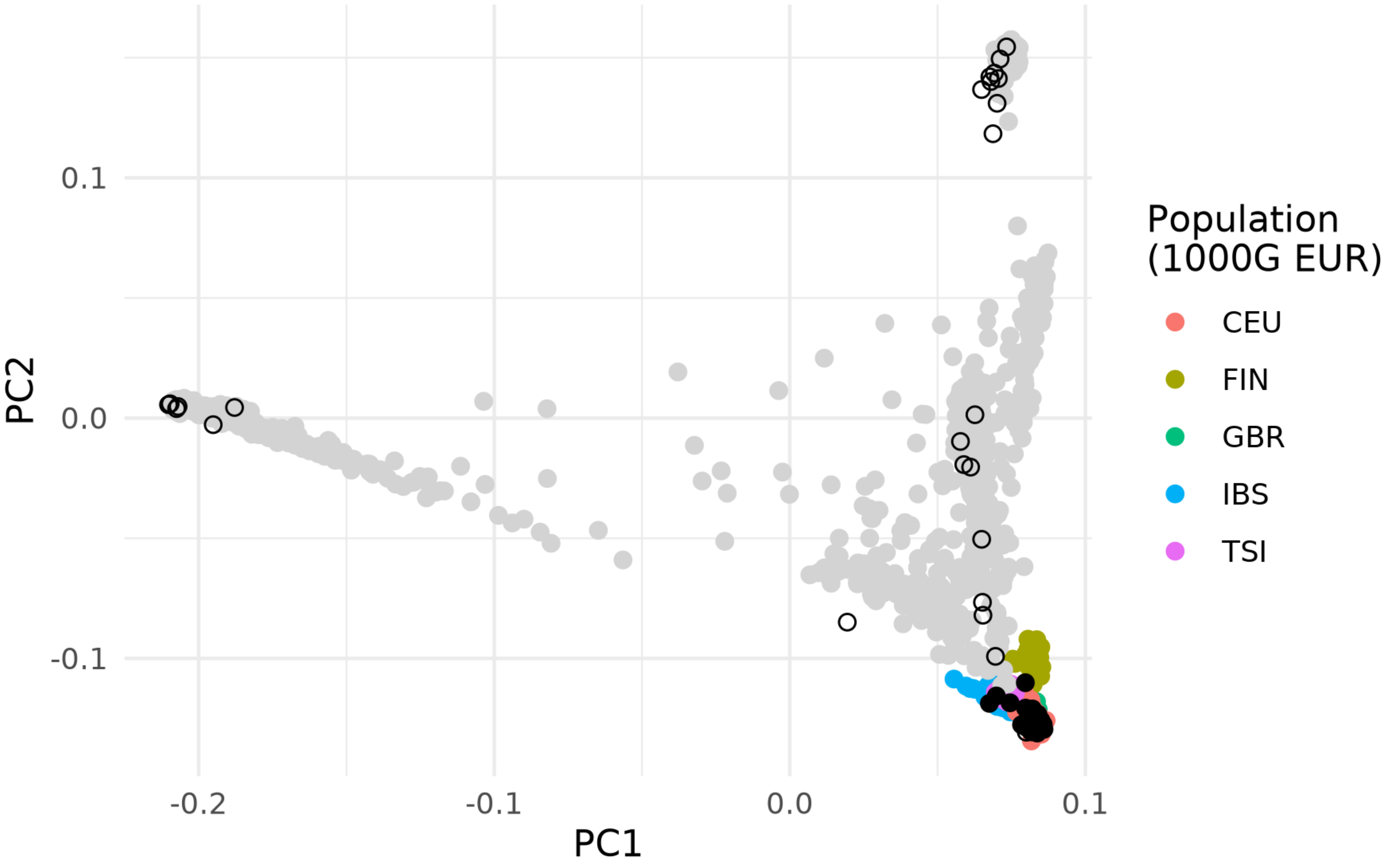


**Supplementary Figure 7:** Principal component analysis of the COMBAT individuals combined with 1000 Genomes (Auton et al., 2015). Individuals from 1000 Genomes European (EUR) populations were highlighted in colour, European−assigned COMBAT individuals showed as filled black circles and non−European samples as non−filled black circles.

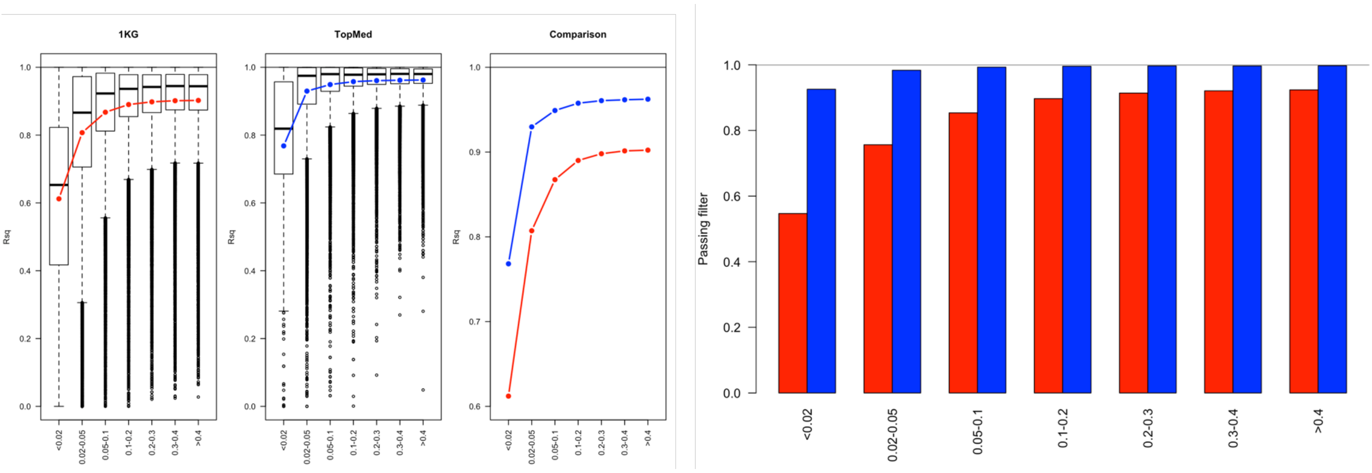


**Supplementary Figure 8:** Imputation quality and proportion of variants passing filters across minor allele frequency bins using 1000 Genomes (1KG) and TOPMed imputation reference sets. Rsq is the squared correlation coefficient between leave−one−one imputation dosage and true genotype dosage for genotyped variants. The filter threshold is predicted R2 > 0.7.

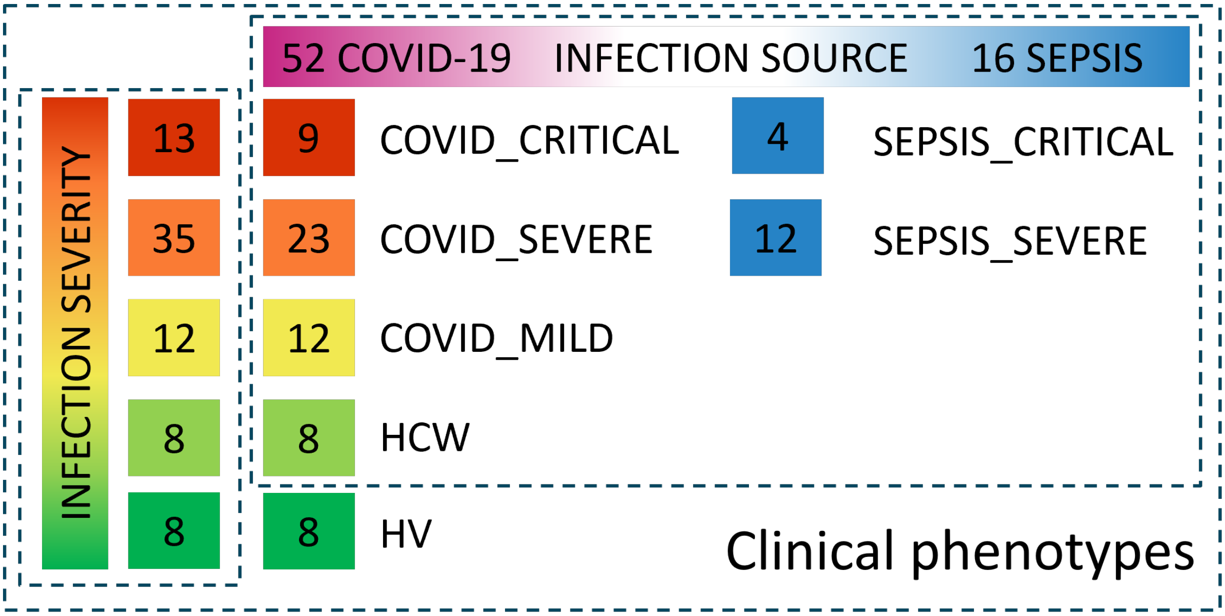


**Supplementary Figure 9:** Categorisation of phenotype groups for disease term in the interaction model. Infection severity comparison includes all individuals, infection source (COVID−19 vs. sepsis) comparison does not include healthy invididuals. When used in association models, infection severity is considered a linear predictor with HV = 1 and CRITICAL = 5, and infection source is considered a binary predictor with COVID−19 = 1 and SEPSIS = 0.

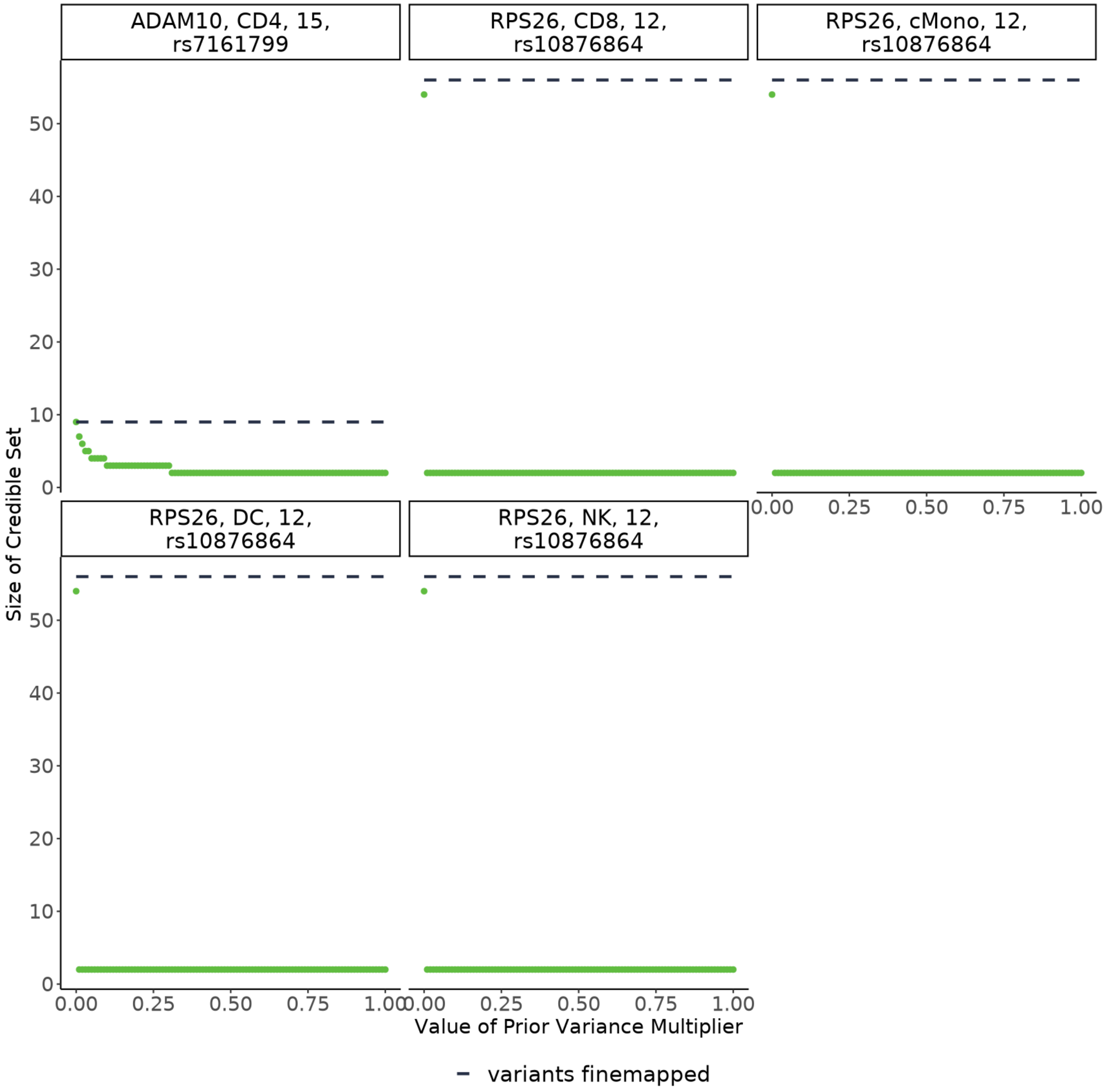


**Supplementary Figure 10:** Sensitivity analysis of the effect of prior variance multiplier on the size of the resultant credible set. The x−axis shows the different values of prior variance multiplier used in the sensitivity analysis. The y−axis shows the size of the credible set after finemapping. In most cases, only values very close 0 affect the result. A value of 0.2 was used in accordance with susieR.
